# Multicenter Invasive Validation of the 2025 ASE Versus 2016 ASE/EACVI Guidelines for the Assessment of Left Ventricular Filling Pressures

**DOI:** 10.1101/2025.09.08.25335376

**Authors:** Amer Barakat, Omar Alkassem, Ahmad Rasheed Alsaadi

## Abstract

**Background:** Accurate assessment of left ventricular filling pressures (LVFP) is critical for the diagnosis and management of diastolic dysfunction. The 2016 ASE/EACVI algorithm has been invaluable in routine practice, yet invasive studies have shown important gaps— especially in sensitivity. The 2025 ASE update brings revised thresholds, integrates and emphasizes left atrial strain, aiming to close these gaps and refine patient care. The clinical value of these changes compared to invasive hemodynamic measurements remains uncertain.

**Objectives:** This study represents an invasive, multicenter validation of the 2025 ASE diastolic function algorithm compared with the long-standing 2016 ASE/EACVI guidelines, using invasive left ventricular end-diastolic pressure (LVEDP) and LV pre-A pressure as the gold standard. Further objectives are to explore subgroup differences based on ejection fraction and sex, as well as, exploring 2-years readmission as an outcome for these guidelines.

**Methods:** Conducting a prospective invasive validation study including patients undergoing comprehensive echocardiography and left heart catheterization. LVEDP ≥ 16 mmHg or LV pre-A > 15 mmHg were defined as elevated filling pressures. Diastolic function was classified according to ASE 2025 and ASE/EACVI 2016 guidelines. Diagnostic accuracy, sensitivity, specificity, and area under the receiver operating characteristic (ROC) curve (AUC) were compared, with subgroup analyses for preserved/reduced EF and sex.

**Results:** A total of 492 patients were included. ASE 2025 demonstrated significantly higher sensitivity than ASE/EACVI 2016 for detecting elevated LVEDP (56.2% vs 22.2%, p<0.00001) and LV pre-A positivity (68.9% vs 25.7%, p<0.00001), while maintaining comparable specificity (for LV pre-A: 82.4%, for LVEDP: 92.6%). ASE 2025 achieved higher AUC values across all subgroups, particularly in preserved EF (0.754 vs 0.577 for LV pre-A) and female patients (0.756 vs 0.576 for LV pre-A). Agreement analysis showed moderate concordance between ASE 2025 and invasive measures (κ=0.46 for LVEDP, κ=0.51 for LV pre-A). In a 2-year follow-up, positive LAP classification by ASE 2025 was associated with higher odds of readmission (OR=3.1, p=0.0341) compared to ASE/EACVI 2016 (OR=2.5, p=0.037).

**Conclusions:** ASE 2025 guidelines provide improved sensitivity and overall diagnostic performance for detecting invasively confirmed elevated LVFP compared to ASE/EACVI 2016, this includes EF and gender subgroups. LAP assessment by ASE 2025 may predict 2-years readmission slightly better than ASE/EACVI 2016.

## Introduction

left ventricular filling pressures (LVFP) is still a major challenge in the diagnosis, prognosis, and therapeutic guidance of heart failure (HF) in its two forms preserved ejection fraction (HFpEF) and reduced ejection fraction (HFrEF) [1,2]. Accurate estimation of LVFP—particularly left ventricular end-diastolic pressure (LVEDP) and pre-atrial contraction (pre-A) wave positivity—is essential for optimizing treatment strategies and risk stratification [3].

Echocardiography has emerged as the best alternative noninvasive method for assessing LVFP trying as much as possible to be comparable to the gold standard invasive method. From its beginning -by integrating conventional, Doppler, tissue Doppler parameters-this domain has undergone a continuous development to improve its diagnostic value, however, these guidelines and algorithms are expert consensus and need always continuous validation to understand and improve the role of echocardiography [4,5].

To a recent date, the 2016 guidelines from the American Society of Echocardiography (ASE) and the European Association of Cardiovascular Imaging (EACVI) were widely adopted. Although it faced non-avoidable criticisms as many studies demonstrated limitation by reduced sensitivity and a relatively high proportion of indeterminate classifications [6].

In 2025, ASE introduced updated recommendations, integrating new echo indices, trying to improve sensitivity and accuracy and reduce indeterminate assessment [7]. These guidelines emphasized new invasive LVFP cutoffs, refined older parameters’ thresholds and integrated LA strain. This update took benefit from latest validation studies on older guidelines and novel echocardiographic indices such as LA strain–volume loops and blood speckle tracking-derived pressure differences have shown stronger alignment with invasive hemodynamics [8–10].

Despite these advancements, the comparative diagnostic performance of ASE 2025 versus ASE 2016 against invasive gold standards remains not widely explored. A very recent study by Lababidi, Naugeh et al. [11] showed promising results, however, this algorithm still need further assessment.

This study addresses this gap by providing first extra-validation (outer from ASE labs) multicenter invasive validation of the new 2025 guidelines compared with the 2016 criteria, incorporating subgroup analyses by ejection fraction (EF) and sex, and examining associations with clinical outcomes such as readmission [7,12].

## Methods

### Study Design and Population

In a prospective observational study of 492 patients were referred for clinically indicated left heart catheterization and comprehensive transthoracic echocardiography at three multicenter Damascus University hospitals between April 2020 and December 2022. The study protocol was approved by the institutional ethics committee (no. 1489, 23.03.2020) and adhered to the Declaration of Helsinki. Written informed consent was obtained from all participants.

Exclusion criteria included Patients who refused to participate to the study, patients with history of atrial fibrillation (AF), ventricular arrhythmia, cardiac arrest or pericardial constriction, moderate to severe valvular heart diseases (including annular classification), valvular prosthesis, left ventricular assist device (LVAD), non-cardiac pulmonary hypertension, congenital heart disease, poor echocardiographic acoustic windows, inability to undergo catheterization, patients with uncontrolled arrhythmias or hemodynamically unstable patients.

Data collection and follow-up were prospective. After publication of the ASE 2025 guidelines, a retrospective re-analysis was performed to compare the diagnostic performance of the 2016 and 2025 algorithms against invasive hemodynamics.

Clinical outcomes included heart failure or other cardiac-related readmission within two years of the baseline evaluation. Structured telephone interviews at 6, 12, and 24 months confirmed hospitalizations and outcomes in the absence of electronic health records (EHR).

### Echocardiographic Assessment

All patients underwent transthoracic echocardiography immediately before catheterization. Experienced echocardiographers obtained standard apical and parasternal views. Measurements included EF by M-mode, transmitral Doppler indices (E and A velocities, E/A ratio, deceleration time), pulmonary vein S/D ratio and Ar-A duration, LA volume index (LAVI), TR velocity, pulmonary artery systolic pressure (PASP), tissue Doppler velocities (septal, lateral, and average e′), and E/e′ (septal, lateral, and average).

LA reservoir strain (LARS) was assessed using vendor-specific speckle-tracking software.

### Guideline-Based Classification

Diastolic function was classified as normal, Grade I–III dysfunction, or indeterminate using both the ASE/EACVI 2016 and ASE 2025 algorithms [6,7]. LA pressure (LAP) was categorized as normal, elevated, or indeterminate according to each algorithm’s criteria.

### Invasive Hemodynamic Measurements

Catheterization was performed blinded to echocardiography. A 6-Fr pigtail catheter was introduced via femoral or radial access. A fluid-filled transducer was balanced before the measurements. Zeroing was standardized at mid-axillary level with verification of fidelity and calibration prior to each measurement and LV pressures were measured at end-expiration over three consecutive cardiac cycles. Elevated LVEDP was defined as ≥16 mmHg and elevated LV pre-A as >15 mmHg [7]. An exploratory analysis considered filling pressures elevated if either LVEDP ≥16 mmHg or LV pre-A >15 mmHg, reflecting that each parameter captures distinct phases of diastolic function (either positive = elevated LVFP while both normal= normal LVFP).

### Study objectives

The primary objective was to compare the diagnostic performance (sensitivity, specificity, PPV, NPV, and accuracy) of ASE 2025 versus ASE/EACVI 2016 in detecting elevated LVEDP and LV pre-A positivity. Secondary objectives included ROC analysis (AUC), agreement with invasive measures (Cohen’s kappa), subgroup analyses (by EF and sex), and prognostic value for two-year readmission.

### Statistical Analysis

Continuous variables were expressed as mean ± SD and compared with Student’s t-test. Categorical variables were presented as counts and percentages, with Chi-square or Fisher’s exact tests for comparisons. ROC curves and AUCs were computed and compared using DeLong’s method. Agreement was assessed by Cohen’s kappa. A two-tailed p <0.05 was considered significant. Analyses were performed using SPSS v27 and R v4.2.

## Results

### Baseline Characteristics

Patients with elevated LV pre-A >15 mmHg (Table 1-Main Tables) had lower lateral e′ (10.35 ± 3.16 vs 10.96 ± 3.07 cm/s, p=0.0345) and higher averaged E/e′ (9.41 ± 4.34 vs 7.85 ± 3.15, p=0.0005), with higher mitral E velocity, E/A ratio, TR velocity, PASP, S/D ratio, Ar-A duration, and lower LA strain (all p<0.0001). Comorbidities such as diabetes (74% vs 50.5%, p<0.0001), ischemic heart disease (60.4% vs 44.3%, p=0.0009), and female sex (51.5% vs 38.4%, p=0.0071) were also more prevalent.

**Table 1:**
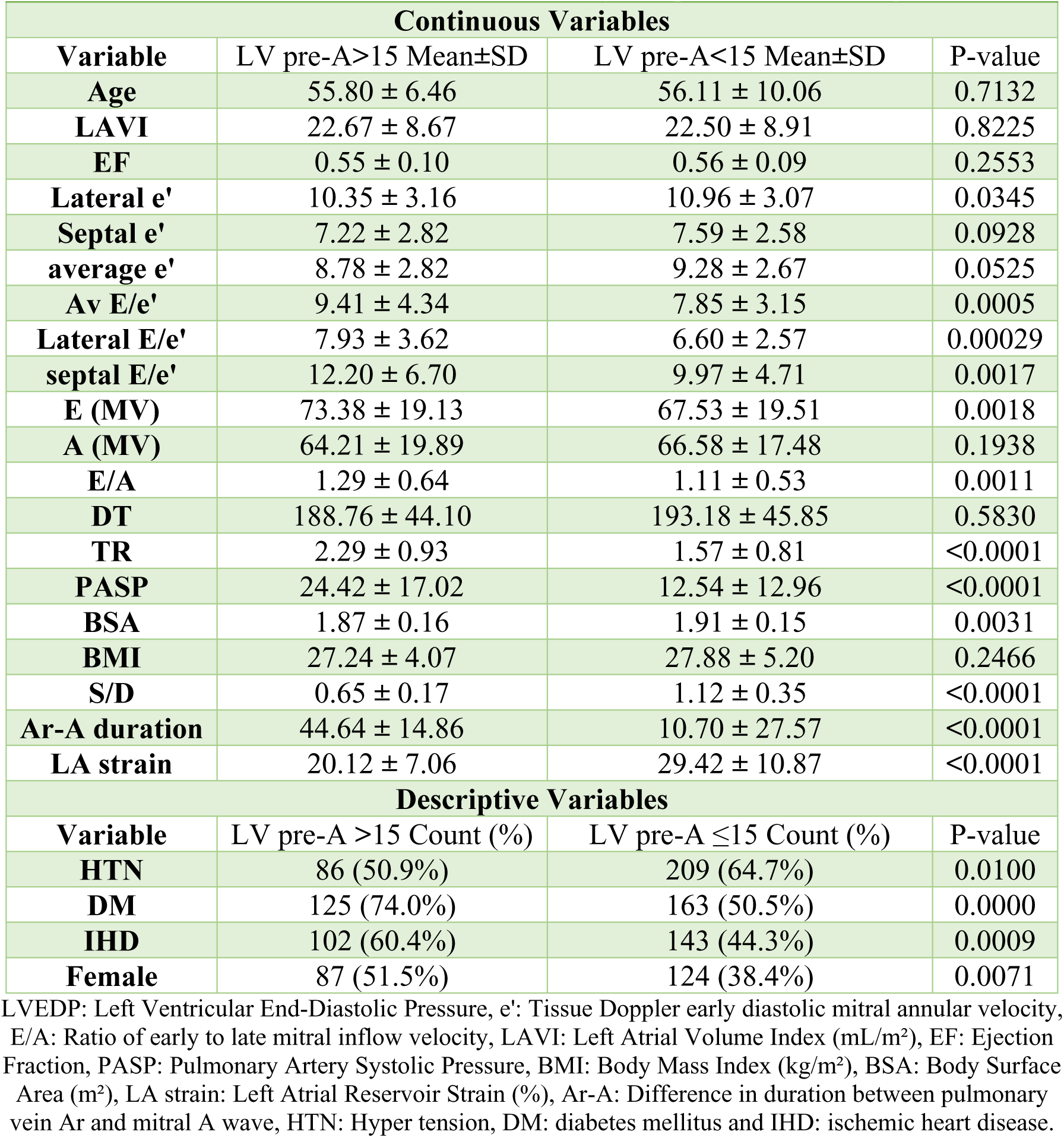
Baseline Characteristics by LV pre-A > 15 mmHg.

| Table 1: Baseline Characteristics by LV pre-A > 15 mmHg |  |  |  |
| --- | --- | --- | --- |
| Continuous Variables |  |  |  |
| Variable | LV pre-A>15 Mean±SD | LV pre-A<15 Mean±SD | P-value |
| Age | 55.80 ± 6.46 | 56.11 ± 10.06 | 0.7132 |
| LAVI | 22.67 ± 8.67 | 22.50 ± 8.91 | 0.8225 |
| EF | 0.55 ± 0.10 | 0.56 ± 0.09 | 0.2553 |
| Lateral e' | 10.35 ± 3.16 | 10.96 ± 3.07 | 0.0345 |
| Septal e' | 7.22 ± 2.82 | 7.59 ± 2.58 | 0.0928 |
| average e' | 8.78 ± 2.82 | 9.28 ± 2.67 | 0.0525 |
| Av E/e' | 9.41 ± 4.34 | 7.85 ± 3.15 | 0.0005 |
| Lateral E/e' | 7.93 ± 3.62 | 6.60 ± 2.57 | 0.00029 |
| septal E/e' | 12.20 ± 6.70 | 9.97 ± 4.71 | 0.0017 |
| E (MV) | 73.38 ± 19.13 | 67.53 ± 19.51 | 0.0018 |
| A (MV) | 64.21 ± 19.89 | 66.58 ± 17.48 | 0.1938 |
| E/A | 1.29 ± 0.64 | 1.11 ± 0.53 | 0.0011 |
| DT | 188.76 ± 44.10 | 193.18 ± 45.85 | 0.5830 |
| TR | 2.29 ± 0.93 | 1.57 ± 0.81 | <0.0001 |
| PASP | 24.42 ± 17.02 | 12.54 ± 12.96 | <0.0001 |
| BSA | 1.87 ± 0.16 | 1.91 ± 0.15 | 0.0031 |
| BMI | 27.24 ± 4.07 | 27.88 ± 5.20 | 0.2466 |
| S/D | 0.65 ± 0.17 | 1.12 ± 0.35 | <0.0001 |
| Ar-A duration | 44.64 ± 14.86 | 10.70 ± 27.57 | <0.0001 |
| LA strain | 20.12 ± 7.06 | 29.42 ± 10.87 | <0.0001 |
| Descriptive Variables |  |  |  |
| Variable | LV pre-A >15 Count (%) | LV pre-A ≤15 Count (%) | P-value |
| HTN | 86 (50.9%) | 209 (64.7%) | 0.0100 |
| DM | 125 (74.0%) | 163 (50.5%) | 0.0000 |
| IHD | 102 (60.4%) | 143 (44.3%) | 0.0009 |
| Female | 87 (51.5%) | 124 (38.4%) | 0.0071 |

LVEDP: Left Ventricular End-Diastolic Pressure, e': Tissue Doppler early diastolic mitral annular velocity, E/A: Ratio of early to late mitral inflow velocity, LAVI: Left Atrial Volume Index (mL/m<sup>2</sup>), EF: Ejection Fraction, PASP: Pulmonary Artery Systolic Pressure, BMI: Body Mass Index (kg/m<sup>2</sup>), BSA: Body Surface Area (m<sup>2</sup>), LA strain: Left Atrial Reservoir Strain (%), Ar-A: Difference in duration between pulmonary vein Ar and mitral A wave, HTN: Hyper tension, DM: diabetes mellitus and IHD: ischemic heart disease.

Elevated LVEDP (≥16 mmHg) was associated with significantly lower EF, reduced septal e′, higher averaged E/e′, higher TR velocity, higher PASP, and markedly reduced LA strain (20.5 ± 6.7% vs 33.9 ± 10.2%, p<0.0001) (Supplementary Table 1).

### Hemodynamic Comparison

No significant differences were observed between echocardiographic and invasive measurements of heart rate, systolic/diastolic blood pressure, or mean arterial pressure (Table 2, Main Tables).

**Table 2:** Hemodynamic Comparison (Echocardiography vs Catheterization)

|  |  |  |  |
| --- | --- | --- | --- |
| Variable | Echo Mean ± SD | Cath Mean ± SD | P-value |
| Heart Rate (bpm) | 72 ± 10 | 74 ± 11 | 0.42 |
| Systolic BP (mmHg) | 130 ± 15 | 132 ± 14 | 0.55 |
| Diastolic BP (mmHg) | 75 ± 10 | 76 ± 9 | 0.61 |
| Mean Arterial Pressure (mmHg) | 93 ± 11 | 95 ± 10 | 0.47 |

Comparison of key hemodynamic parameters measured during echocardiography vs cardiac catheterization. Values are presented as Mean ± SD.

### Clinical History and Medications

Table 3 (Main Tables) summarizes medical history and medication use. Dyspnea (NYHA ≥ II) was reported in 66.7% of patients, orthopnea in 22.5%, and fatigue in 34.8%. Common comorbidities included chronic kidney disease (20.3%), hypertension and diabetes mellitus.

**Table 3:** Clinical History, Medications, and Symptoms.

| Item | Status | percentage (%) |
| --- | --- | --- |
| **Chronic Kidney Disease (eGFR <60)** | Yes | (20.3%) |
| **Beta-blockers** | On medication | (64.5%) |
| **ACEi/ARB** | On medication | (58.7%) |
| **Diuretics** | On medication | (43.5%) |
| **Statins** | On medication | (52.9%) |
| **Dyspnea (NYHA ≥ II)** | Present | (66.7%) |
| **Orthopnea** | Present | (22.5%) |
| **Fatigue** | Present | (34.8%) |

Medical history and symptom data presented as number of patients and corresponding percentage. ACEi = Angiotensin-Converting Enzyme inhibitor; ARB = Angiotensin II Receptor Blocker; NYHA = New York Heart Association functional class.

### LAP Classification

ASE 2025 identified more patients with elevated LAP (34.6% vs 14.6%, p<0.0001) and fewer indeterminate cases (2.6% vs 8.7%, p<0.0001) compared with ASE 2016 (Table 4, Main Tables).

**Table 4:** Comparison between ASE 2025 and ASE/EACVI 2016 for the assessment of LAP.

| Algorithm | Normal LAP Count (%) | Elevated LAP Count (%) | Indeterminate LAP Count (%) |
| --- | --- | --- | --- |
| **ASE 2025** | 309 (62.8%) | 170 (34.55%) | 13 (2.64%) |
| **ASE/EACVI 2016** | 377 (76.63%) | 72 (14.63%) | 43 (8.74%) |
| **P value** | <0.0001 | <0.0001 | <0.0001 |

ASE: American Society of Cardiology, EACVI: European Association of Cardiovascular Imaging, LAP: left atrial pressure.

### Diagnostic Performance

Across all invasive standards, ASE 2025 outperformed ASE 2016 (Table 5, Main Tables). For LVEDP ≥16 mmHg, sensitivity was 56.2% vs 22.2% (p<0.00001) with comparable specificity (92.6% vs 91.9%). For LV pre-A >15 mmHg, sensitivity was 68.9% vs 25.7%, specificity 82.4% vs 88.5%. Accuracy and ROC AUC values were consistently higher with ASE 2025 (Figures 1–3, Main Figures).

**Figure 1:**
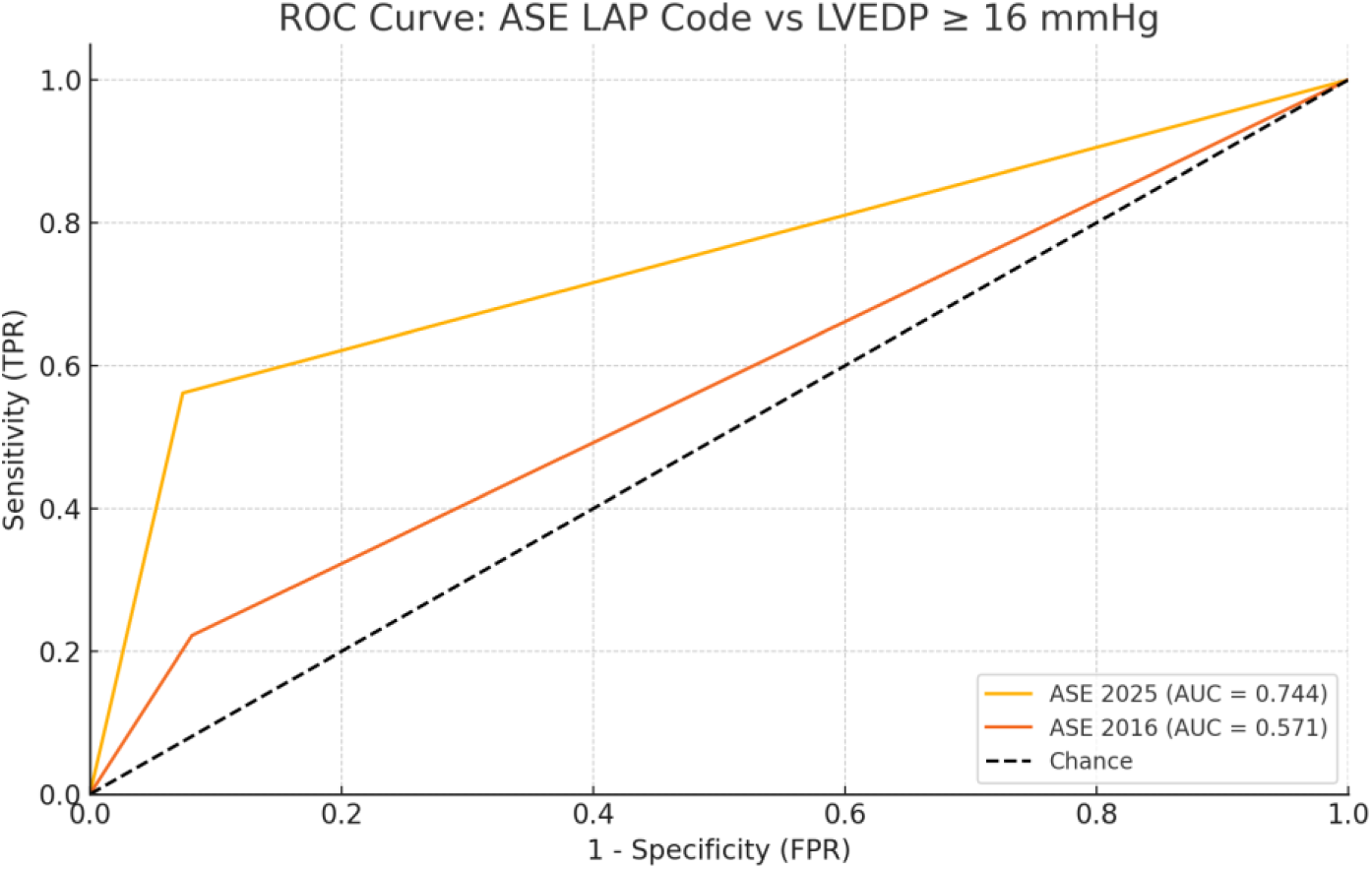
ROC Curve comparing ASE 2025 vs ASE 2016 based on LAP Code classification against invasive LVEDP ≥ 16 mmHg.

**Figure 2:**
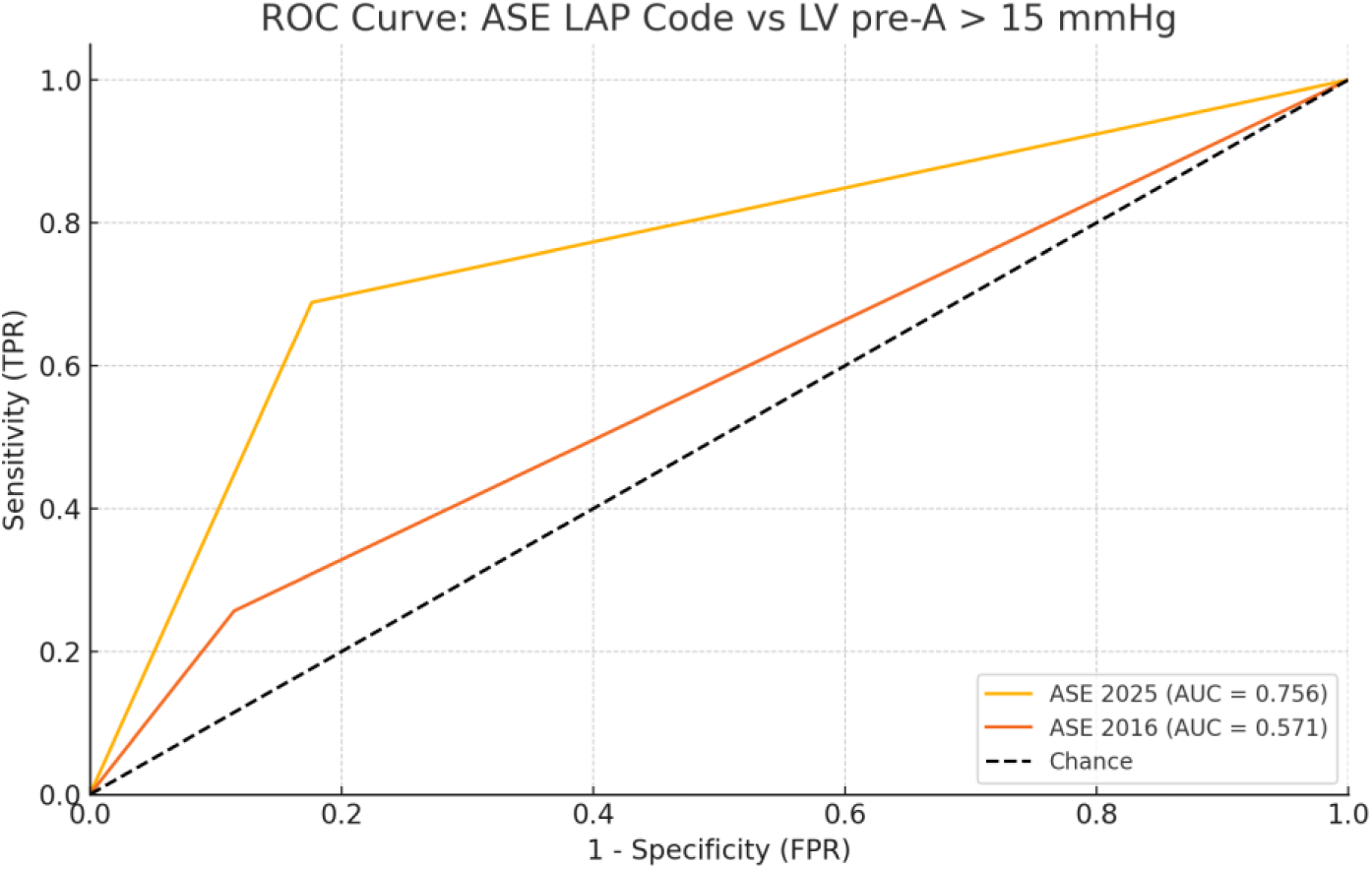
ROC Curve comparing ASE 2025 vs ASE 2016 based on LAP Code classification against invasive LV pre-A > 15 mmHg.

**Figure 3:**
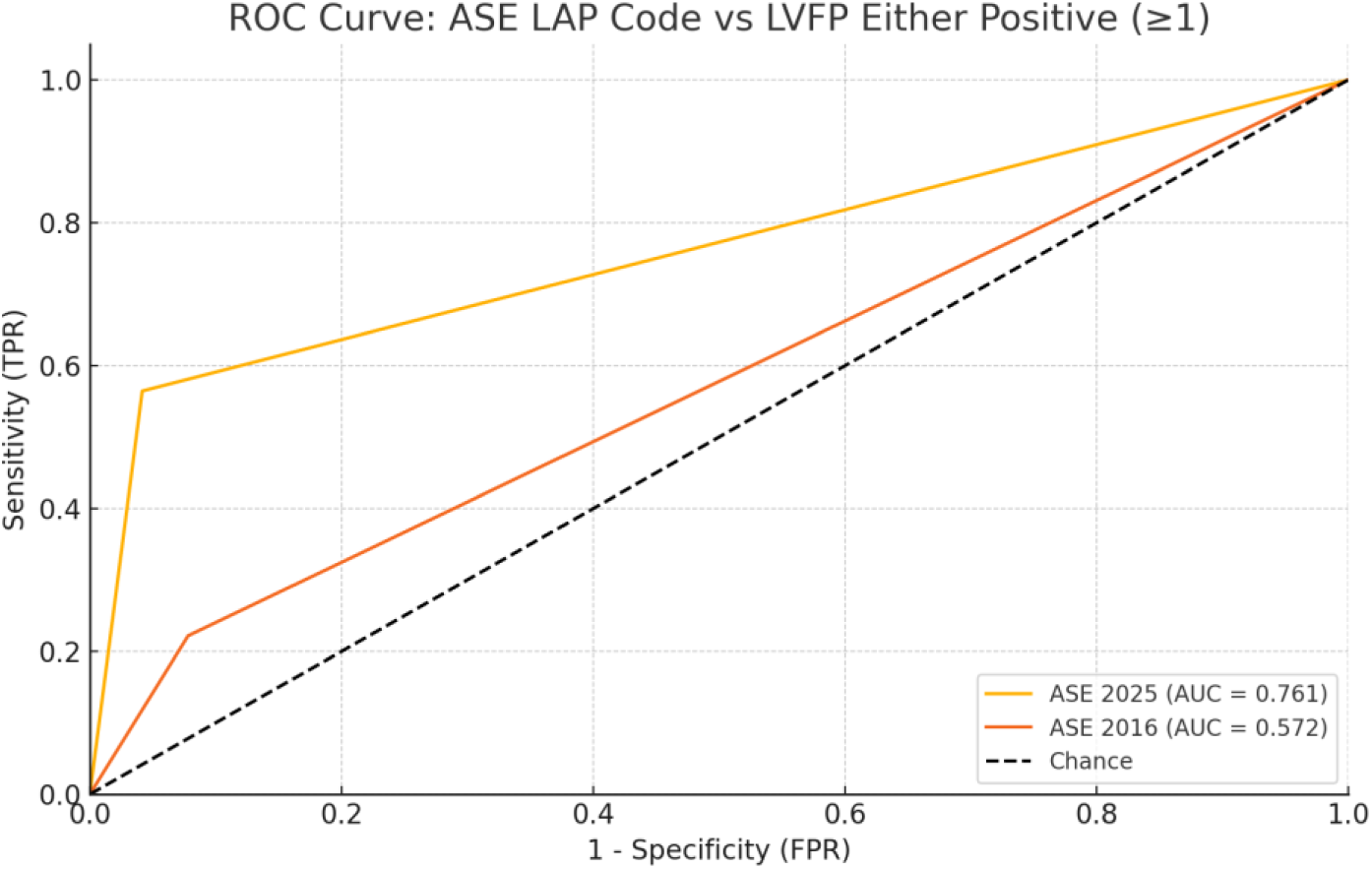
ROC Curve comparing ASE 2025 vs ASE 2016 based on LAP Code classification against invasive LVFP either positive (≥1).

**Figure 4:**
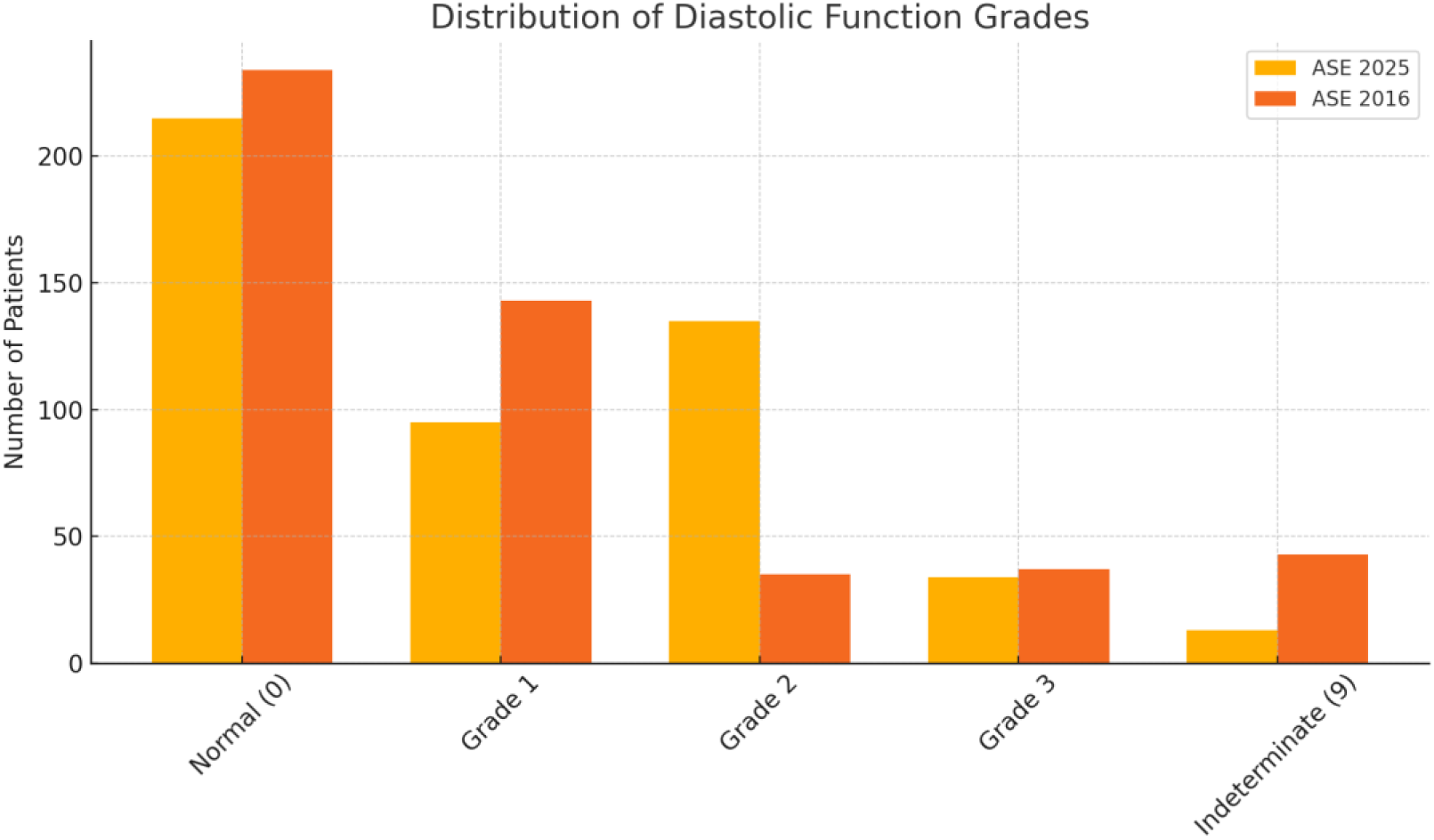
Distribution of diastolic function grades according to ASE 2025 and ASE 2016.

**Table 5:** Comparison between ASE 2025 and ASE/EACVI 2016 diagnostic performance vs different LVFP waves.

| LV pressure wave | Algorithm | Sensitivity | Specificity | PPV | NPV | Accuracy |
| --- | --- | --- | --- | --- | --- | --- |
| **LVEDP ≥ 16 mmHg** | ASE 2025 | 0.562 | 0.926 | 0.912 | 0.608 | 0.716 |
|  | ASE 2016 | 0.222 | 0.919 | 0.778 | 0.48 | 0.528 |
| **LV pre-A > 15 mmHg** | ASE 2025 | 0.689 | 0.824 | 0.676 | 0.832 | 0.777 |
|  | ASE 2016 | 0.257 | 0.885 | 0.514 | 0.716 | 0.684 |
| **LVFP either positive** | ASE 2025 | 0.564 | 0.958 | 0.953 | 0.595 | 0.722 |
|  | ASE 2016 | 0.222 | 0.922 | 0.792 | 0.469 | 0.521 |

ASE: American Society of Cardiology, EACVI: European Association of Cardiovascular Imaging,
LVEDP: left ventricular end diastolic pressure, LVFP: left ventricular filling pressure, P-value for difference in Sensitivity between ASE 2025 and ASE 2016: <0.00001 and Statistical test used: Chi-square test for 2x2 contingency table.

### Readmission Outcomes

ASE 2025 LAP positivity predicted two-year readmission more strongly (OR=3.1, 95% CI 2.91–3.37, p=0.0341) than ASE 2016 (OR=2.5, p=0.037) (Table 6, Main Tables).

**Table 6:** comparison of further admission or re-hospitalization in two years.

|  |  |  |  |  |
| --- | --- | --- | --- | --- |
| **Variable** | **OR** | **95% CI Lower** | **95% CI Upper** | **P-value** |
| **ASE 2025 LAP positive** | 3.1 | 2.906 | 3.371 | 0.0341 |
| **ASE/EACVI 2016 LAP positive** | 2.5 | 1.942 | 2.95 | 0.0370 |

ASE: American Society of Cardiology, EACVI: European Association of Cardiovascular Imaging, LAP: left atrial pressure.

### Diastolic Function Grading

ASE 2025 classified more patients as Grade II dysfunction (27.4% vs 7.5%) and fewer as indeterminate (Table 7, Main Tables). Agreement analysis showed moderate concordance with invasive measures (κ=0.456 for LVEDP, κ=0.51 for LV pre-A) (Supplementary Table 2).

**Table 7:** Diastolic grade distribution between ASE 2025 and ASE/EACVOI 2016.

|  |  |  |  |  |  |
| --- | --- | --- | --- | --- | --- |
| **Algorithm** | **Normal** | **Grade 1** | **Grade 2** | **Grade 3** | **Indeterminate** |
| **ASE 2025** | 215<br>(43.7%) | 95<br>(19.3%) | 135 (27.4%) | 34 (6.9%) | 13 (2.6%) |
| **ASE/EAVI 2016** | 234<br>(47.6%) | 143<br>(29.1%) | 35 (35%) | 37 (7.1%) | 43 (7.5%) |
| **P value** | <0.0001 | <0.0001 | <0.0001 | <0.0001 | <0.0001 |

ASE: American Society of Cardiology, EACVI: European Association of Cardiovascular Imaging, P-values calculated using Chi-square test

### Subgroup Analyses

ASE 2025 achieved higher AUC values across EF and sex subgroups. In preserved EF, AUC for LV pre-A was 0.754 vs 0.577 with ASE 2016. Among women, ASE 2025 reached 0.756 vs 0.576 with ASE 2016 (Table 8, Main Tables; Figures 5–6, Main Figures; Figures 1–3, Supplementary Figures).

**Figure 5:**
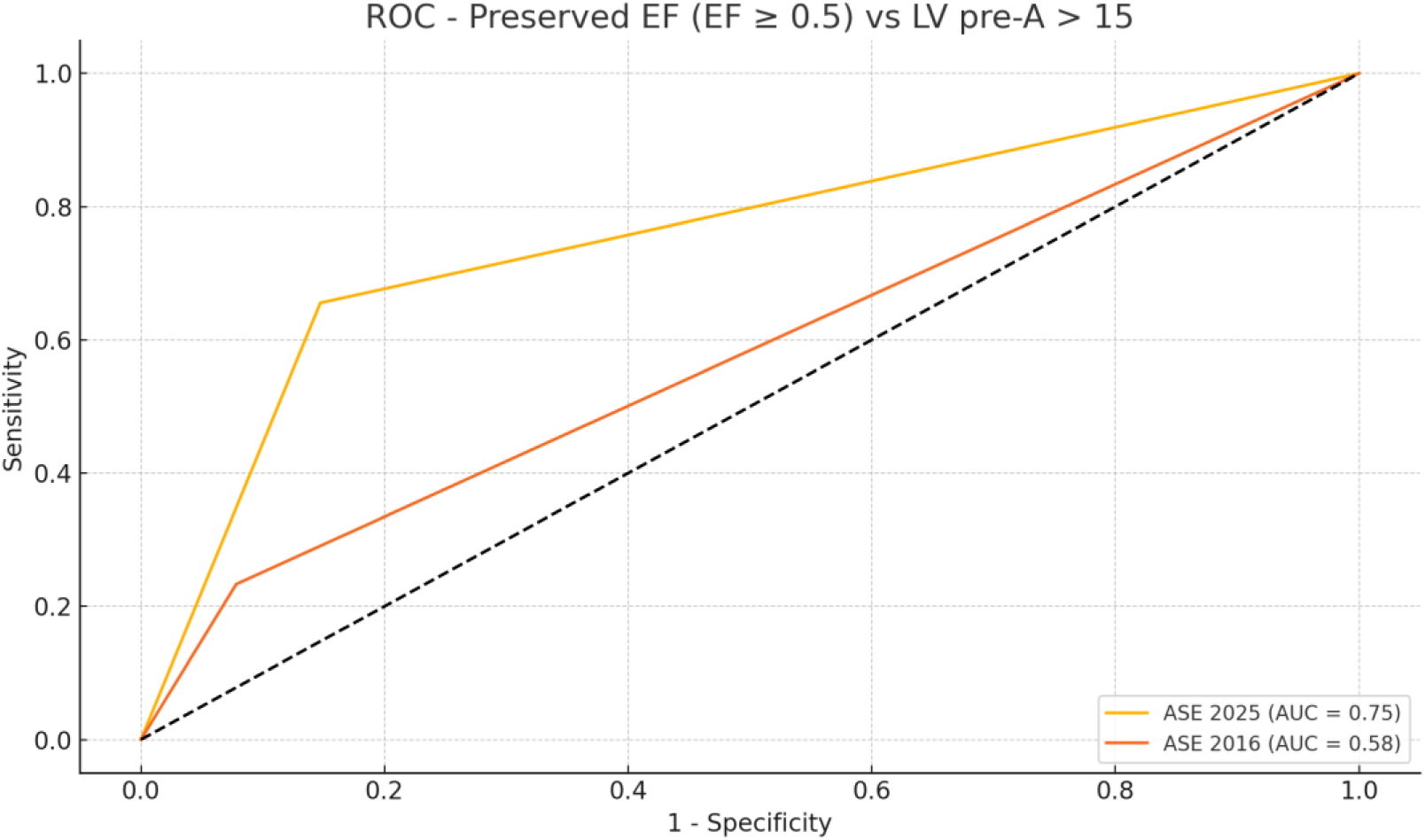
Comparison the diagnostic performance (ROC AUC) of ASE 2025 and ASE 2016 LAP classifications within preserved EF subgroups, using LV pre-A > 15 mmHg.

**Figure 6:**
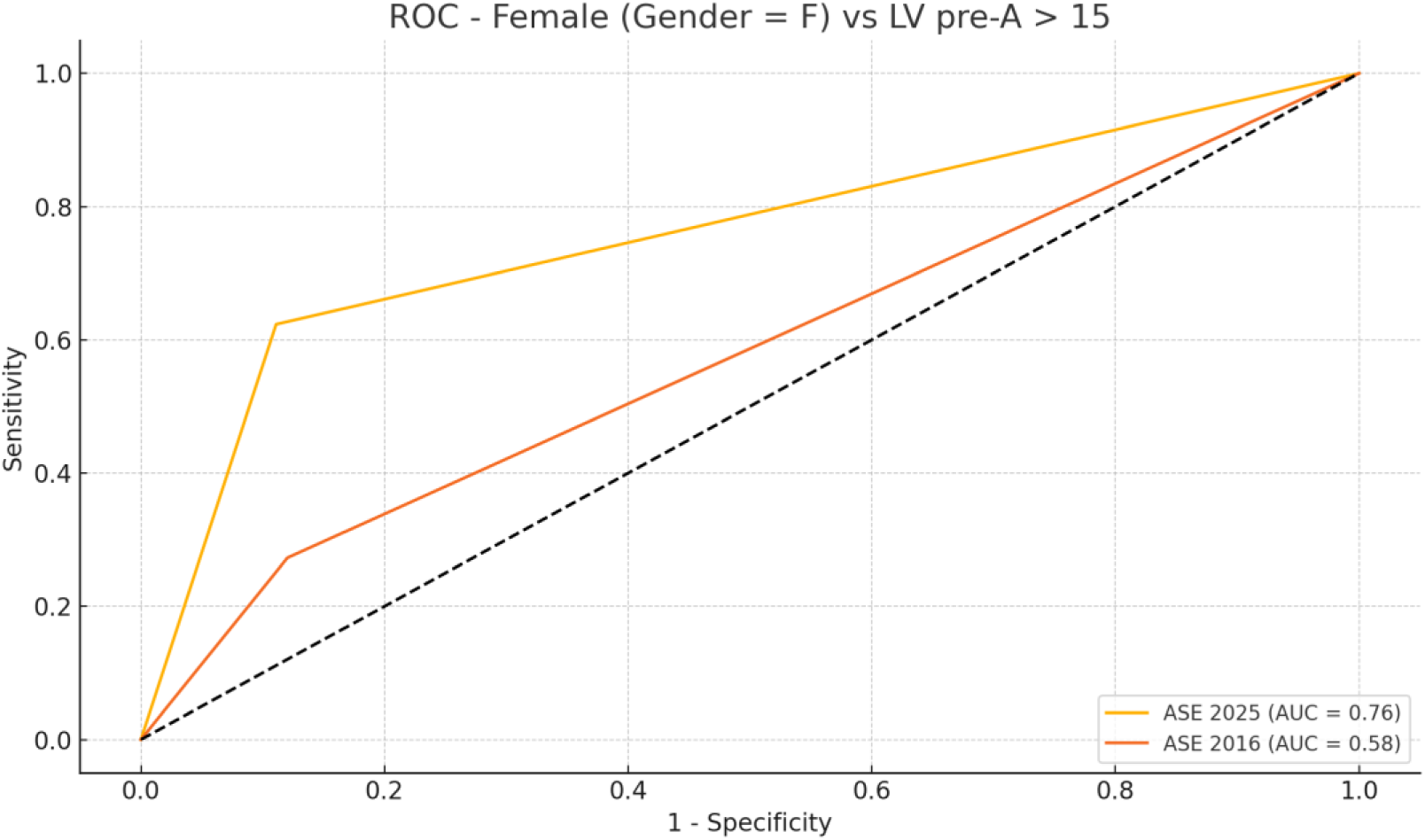
Comparison the diagnostic performance (ROC AUC) of ASE 2025 and ASE 2016 LAP classifications within female subgroups, using LV pre-A > 15 mmHg.

**Table 8:** ROC/AUC comparison of ASE 2025 and ASE/EACVI 2016 LAP classifications within EF subgroups.

|  |  |  |  |
| --- | --- | --- | --- |
| **LV pressure wave** | **EF Group** | **Guideline** | **AUC** |
| **LVEDP ≥ 16 mmHg** | Preserved EF (≥0.5) | ASE 2025 | 0.737 |
| | Preserved EF (≥0.5) | ASE 2016 | 0.572 |
| | Reduced EF ($<$0.5) | ASE 2025 | 0.744 |
| | Reduced EF ($<$0.5) | ASE 2016 | 0.537 |
| **LV pre-A $>$ 15 mmHg** | Preserved EF (≥0.5) | ASE 2025 | 0.754 |
| | Preserved EF (≥0.5) | ASE 2016 | 0.577 |
| | Reduced EF ($<$0.5) | ASE 2025 | 0.721 |
| | Reduced EF ($<$0.5) | ASE 2016 | 0.531 |

ASE: American Society of Cardiology, EACVI: European Association of Cardiovascular Imaging, LVEDP: left ventricular end diastolic pressure, AUC: area under the curve.

### Additional Findings

Comparisons of adjacent diastolic grades with invasive pressures (Supplementary Table 3) revealed that ASE 2025 demonstrated significant pressure separation between Grade 1 and Grade 2 for both LVEDP and LV pre-A, whereas ASE/EACVI 2016 showed less consistent separation. EF subgroup LAP comparisons (Supplementary Tables 4–5) indicated significant classification differences for preserved EF under ASE/EACVI 2016 but not ASE 2025.

## Discussion

This study provides a multicenter invasive validation of the ASE 2025 guidelines, demonstrating superior diagnostic accuracy, fewer indeterminate cases, and stronger prognostic value compared with the 2016 criteria.

In this prospective invasive validation study, the updated 2025 ASE guidelines for evaluating left ventricular filling pressure (LVFP) demonstrated substantially higher diagnostic sensitivity compared to the 2016 criteria, while maintaining high specificity. The 2025 algorithm resulted in markedly fewer indeterminate classifications (Table 4), elevated AUC in ROC analysis across the entire cohort (Figures 1–3), and stronger correlations with two-year readmission risk (Table 6). These findings underscore the robustness and clinical relevance of the new guideline.

Baseline echocardiographic parameters stratified by LV pre-A > 15 mmHg (Table 1) and by LVEDP ≥ 16 mmHg (Supplementary Table 1) revealed significant differences in tissue Doppler indices, TR velocity, LA strain, and comorbidities such as diabetes mellitus and ischemic heart disease. These invasive-validated differences confirm that the ASE 2025 –as it has better diagnostic performance than ASE/EACVI 2016-algorithm integrates parameters more tightly linked with true hemodynamic burden.

Hemodynamic comparison between echocardiography and catheterization (Table 2) demonstrated no significant differences in heart rate, blood pressure, or mean arterial pressure, validating the internal consistency and non-bias of our echo dataset relative to invasive standards.

Clinical history, comorbidities, and medical therapy (Table 3) illustrate that the study cohort represented a typical high-risk population, with substantial prevalence of HF symptoms (NYHA ≥ II), CKD, hypertension, and diabetes. These characteristics further highlight the clinical relevance of accurate noninvasive LVFP assessment.

Comparison of LAP classifications (Table 4) shows ASE 2025 identified a higher proportion of patients with elevated LAP and dramatically reduced the proportion of indeterminate cases. This feature improves diagnostic clarity in daily practice.

Diagnostic performance metrics (Table 5) confirmed that ASE 2025 significantly outperformed ASE 2016 for both LVEDP ≥ 16 mmHg and LV pre-A > 15 mmHg. The ROC curves (Figures 1–3) graphically illustrate these superior discriminations.

An exploratory analysis -LVFP either wave positive-was also considered as in definition of invasively elevated filling pressure only one wave is sufficient to establish the diagnose invasively, while this is not how the algorithms work as they oriented to LAP. However, ASE 2025 showed a good diagnostic performance with high specificity and moderate sensitivity for this exploratory analysis. This expanded invasive definition acknowledges that LVEDP and LV pre-A reflect different aspects of diastolic filling, and either may be abnormal in patients with HFpEF. Although not mandated by ASE guidelines, this approach may provide a more sensitive reference standard.

ROC/AUC subgroup analyses (Table 8, Figures 5–6, Supplementary Figures 1–6) confirmed the improved diagnostic accuracy across preserved and reduced EF populations as well as between male and female patients. In preserved EF in particular, ASE 2025 reached an AUC of 0.754 for LV pre-A compared with only 0.577 for ASE 2016.

Readmission analysis (Table 6) revealed that ASE 2025 LAP positivity carried a higher odds ratio for re-hospitalization compared with ASE 2016, underscoring stronger prognostic implications.

Similarly, distribution of diastolic grades (Table 7, Figure 4, Figure 7) demonstrated that ASE 2025 reclassified a greater proportion into Grade 2, reducing indeterminate grading and aligning better with invasive pressure differences (Supplementary Table 3), especially the converting grades between one and two which has thhe overlap between normal and elevated filling pressure assessment.

**Figure 7:**
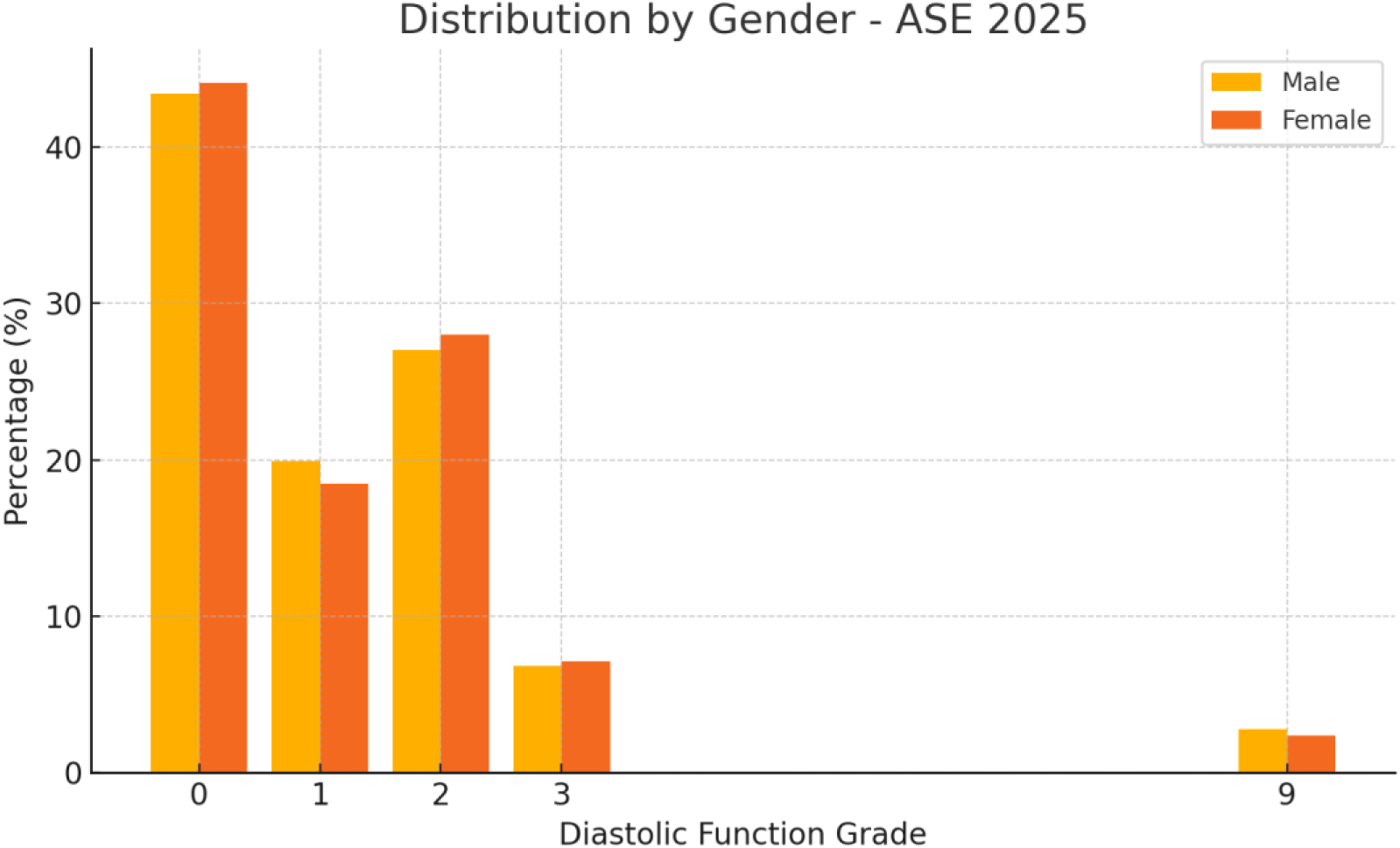
diastolic function distribution according to ASE 2025 between Gender subgroups, 0: normal, 1: Grade 1, 2: Grade 2, 3: Grade 3, 9: indeterminate.

Agreement analysis (Supplementary Table 2) confirmed moderate concordance of ASE 2025 with invasive standards, while subgroup LAP comparisons by EF (Supplementary Tables 4–5) and gender-based distributions (Supplementary Table 6, Supplementary Figure 7) reinforced consistency of the 2025 algorithm across diverse patient profiles.

These findings align with global literature: many studies investigated the diagnostic accuracy of 2016 algorithm, Andersen et al. [3] showed a sensitivity=0.87 and specificity= 0.88. Balaney et al. [13] showed a sensitivity=0.69 and specificity= 0.81. obokata et al. [14] showed a sensitivity=0.34 and specificity= 0.83. Lancelloti et al [15] showed a sensitivity=0.75 and specificity= 0.74. and Sato et al. [16] showed a sensitivity= 0.34 and specificity= 0.86.

These results reflect the widely variation of the sensitivity of the 2016 guidelines (pooling together sensitivity of 61%) [17] among these studies, which raised the questions about the real sensitivity from a hand, and the possible ways to improve this accuracy from another hand.

A relatively recent study in September 2021, Arno A. van de Bovenkamp et al. [18] showed The correlation between diastolic function grades of the ASE/EACVI algorithm and PCWP was poor (r=0.17, P<0.05). The ASE/EACVI algorithm had a sensitivity and specificity of 35% and 87%, respectively; an accuracy of 67% and an area under the curve of 0.56. Moreover, in 30% of cases the algorithm was not applicable or indeterminate.

Due to lack of validation of the very recent ASE 2025 guidelines till now, a literature overview of the new echocardiographic technique to discuss the benefit of these methods would be beneficial: Sørensen et al. (2025) [9] validated intraventricular pressure difference (IVPD) with blood speckle tracking (BST) and concluded that BST echocardiography provides accurate estimation of IVPD in early diastole; Venkateshvaran et al. (2024) [8] emphasized the role of (LASr) and LA strain-volume loops over conventional echocardiographic measures in the identification of elevated LV filling pressure; The PROMIS-HFpEF study highlighted atrial dysfunction’s role in HFpEF [19]. ASE 2025 update itself integrates advanced imaging and novel computational models suggest future integration of strain-based simulations [7].

Lababidi, Naugeh et al. [11] showed that new algorithm increases the feasibility of estimating LVFP and has good accuracy (sensitivity was 86% and specificity was 86%, with accuracy of 86%.) and Only 2 patients had indeterminate LVFP as per the new algorithm versus 38 applying 2016 guidelines (P<0.0001) in 949 cohort. These results come along with the results of this study [ASE 2025 identified fewer indeterminate cases (2.6% vs 8.7%, p<0.0001) compared with ASE 2016] (Table 4, Main Tables). Similar diagnostic results Across all invasive standards, ASE 2025 outperformed ASE 2016 (Table 5, Main Tables) with modest sensitivity for LVEDP ≥16 mmHg, 56.2% vs 22.2% (p<0.00001) and for LV pre-A >15 mmHg, sensitivity was 68.9% vs 25.7%.

This external invasive validation of guideline explored the prognostic role and gender subgroups making the current findings uniquely valuable.

## Conclusion

The ASE 2025 diastolic function guidelines demonstrate superior diagnostic sensitivity for elevated LVEDP and LV pre-A positivity without compromising specificity, compared with ASE/EACVI 2016. The updated algorithm reduces indeterminate cases, improves grade discrimination, and shows robust performance across EF and sex subgroups. These findings support the integration of ASE 2025 into routine clinical practice and HFpEF assessment.

Future research should integrate ASE 2025 criteria with advanced imaging and multimodality validation to refine accuracy and prognostic value.

## Limitations

- Echocardiography and catheterization were not simultaneous, though performed immediately before cath without statistically difference between hemodynamics to minimize the error.
- Referral bias may exist, as patients undergoing invasive evaluation represent a higher-risk cohort.
- Outcomes beyond two years were not assessed.
- Advanced echocardiographic parameters (e.g., GLS, 3D) were not included.
- Exploratory analyses using combined invasive definitions are not echo-guideline-based and should be interpreted cautiously.

## Data Availability Statement

De-identified study data, including echocardiographic measurements and invasive hemodynamics, are available from the corresponding author upon reasonable request, compliant with ethical approvals and data-use agreements.

